# Pre-procedural testing using patient-specific models is associated with high training fidelity and improved procedural efficiency in endovascular aneurysm treatment

**DOI:** 10.64898/2026.04.23.26351592

**Authors:** Jeremy Hofmeister, Gianmarco Bernava, Andrea Rosi, Olivier Brina, Philippe Reymond, Michel Muster, Karl-Olof Lovblad, Paolo Machi

## Abstract

**Background:** Even for experienced operators, endovascular treatment of unruptured intracranial aneurysms involves intraoperative uncertainty that may lead to adjustments in strategy, prolong the procedure, and potentially cause inefficiency and device waste. This study aimed to evaluate whether pre-procedural testing (PPT) of endovascular treatment using patient-specific models was associated with increased operator confidence and perceived clinical utility, including improvements in procedural efficiency and reduced resource waste.

**Methods:** We enrolled a cohort of patients who underwent PPT before endovascular treatment for complex unruptured intracranial aneurysms and compared their outcomes with a control group treated without PPT. The primary outcome was the Training Fidelity Score, a composite of three operator-reported Likert items defined a priori. Secondary outcomes included perceived clinical utility, intraoperative strategy changes, procedural time, radiation exposure, device waste and safety.

**Results:** A total of 85 patients met the inclusion criteria (PPT=40; control=45). The Training Fidelity Score was high across the PPT group (median, 4.33/5). Perceived clinical utility was high and further increased significantly after the procedure. A significant reduction was observed in intraoperative strategy changes, with no changes recorded in the PPT group, compared to 6/45 in the control group (RR 0.09; p=0.027). Reductions in treatment time, radiation exposure and device waste were also noted.

**Conclusion:** PPT using patient-specific models was associated with increased operator confidence, fewer intraoperative strategy changes, improved procedural efficiency, and reduced device waste without compromising safety. These findings support its use in pre-interventional preparation, but require prospective multicenter validation.

## Introduction

Endovascular treatment of unruptured intracranial aneurysms has evolved substantially over the past decades, with improvements in device technology and imaging quality.[1–4] Despite these advances, procedural variability remains inherent to aneurysm treatment. Even for experienced operators, intra-procedural adjustments are common and may prolong procedural time, increase radiation exposure, and lead to device exchanges or waste.[5–7] While technical success and angiographic occlusion have traditionally been emphasized, procedural efficiency and resource utilization have received comparatively less attention.

Pre-procedural planning for the endovascular treatment of unruptured intracranial aneurysms currently relies primarily on imaging analysis and cognitive simulation of the anticipated steps. However, this approach does not fully resolve the uncertainty inherent to aneurysm treatment. Even with careful pre-procedural preparation, operators may need to adjust their strategy during the procedure due to discrepancies between the anticipated and actual vascular anatomy or device behavior. Although virtual reality simulators and 3D models have been used for educational and device evaluation in neurointerventional practice, there is limited evidence quantifying whether patient-specific pre-procedural testing (PPT) translates into measurable improvements in procedural performance in routine clinical practice.[8,9] In particular, the impact of such testing on operator confidence in the planned strategy and on procedural duration, radiation exposure and device waste has not been systematically evaluated in a real-world cohort of unruptured intracranial aneurysm treatments.

Conceptually, testing the treatment on a patient-specific benchtop model before the procedure may reduce uncertainty before patient exposure.[10–14] By refining intracranial artery navigation, embolization strategy and device selection, patient-specific testing could strengthen operator confidence in the planned strategy and reduce inefficiencies related to device exchanges, repositioning and prolonged fluoroscopy. Studies in the broader endovascular field have demonstrated that patient-specific pre-procedural rehearsal reduces procedural errors and operative time.[15–18] Initial experiences in the neurointerventional field using patient-specific, 3D-printed aneurysm models have shown preliminary feasibility.[19–23] Therefore, we conducted this single-center, prospective cohort study to evaluate whether PPT using patient-specific models is associated with high operator-reported training fidelity and perceived clinical utility.

## Methods

### Study design and population

We analyzed a cohort of consecutive adult patients (≥18 years) who underwent endovascular treatment for a solitary unruptured intracranial aneurysm at our center to evaluate the perceived utility of PPT and its contribution to procedural outcomes. Patients were categorized into two groups, ie, those who underwent PPT prior to treatment (PPT group) and those who did not (control group).

PPT was reserved for cases that the primary operator considered to be anatomically or technically challenging. Straightforward cases were not considered for PPT. All unruptured intracranial aneurysm cases were initially evaluated by cerebral angiography in accordance with our institutional protocol. After a subsequent multidisciplinary board decision following the angiography, they were treated by experienced, attending neurointerventionalists. All procedures were performed under general anesthesia using a biplane angiography system (Azurion 7 B20/15, Philips Healthcare, Best, The Netherlands). According to institutional protocol, patients received dual antiplatelet therapy before treatment and intraprocedural anticoagulation. A dedicated research assistant prospectively recorded procedural and clinical variables for both groups during and immediately after each intervention. This study was approved by our local ethical committee and is reported in accordance with the Strengthening the Reporting of Observational studies in Epidemiology (STROBE) statement.

### Patient selection

Consecutive adult patients (age ≥18 years) treated for an unruptured intracranial aneurysm at the Interventional Neuroradiology Unit of the Geneva University Hospitals were eligible for inclusion in the study. All included patients underwent pre-procedural 3D rotational angiography. This imaging technique is necessary for fabricating patient-specific models in the PPT group. It also served as the standard reference for procedural planning in both groups. Patients were planned to be excluded from the study if pre-procedural 3D rotational angiography was unavailable or if the procedural data necessary to compute the primary and secondary endpoints was incomplete. All treated aneurysms were unruptured and no patient underwent treatment for multiple aneurysms during the same procedure. A patient selection flow chart is provided in figure 1.

**Figure 1:**
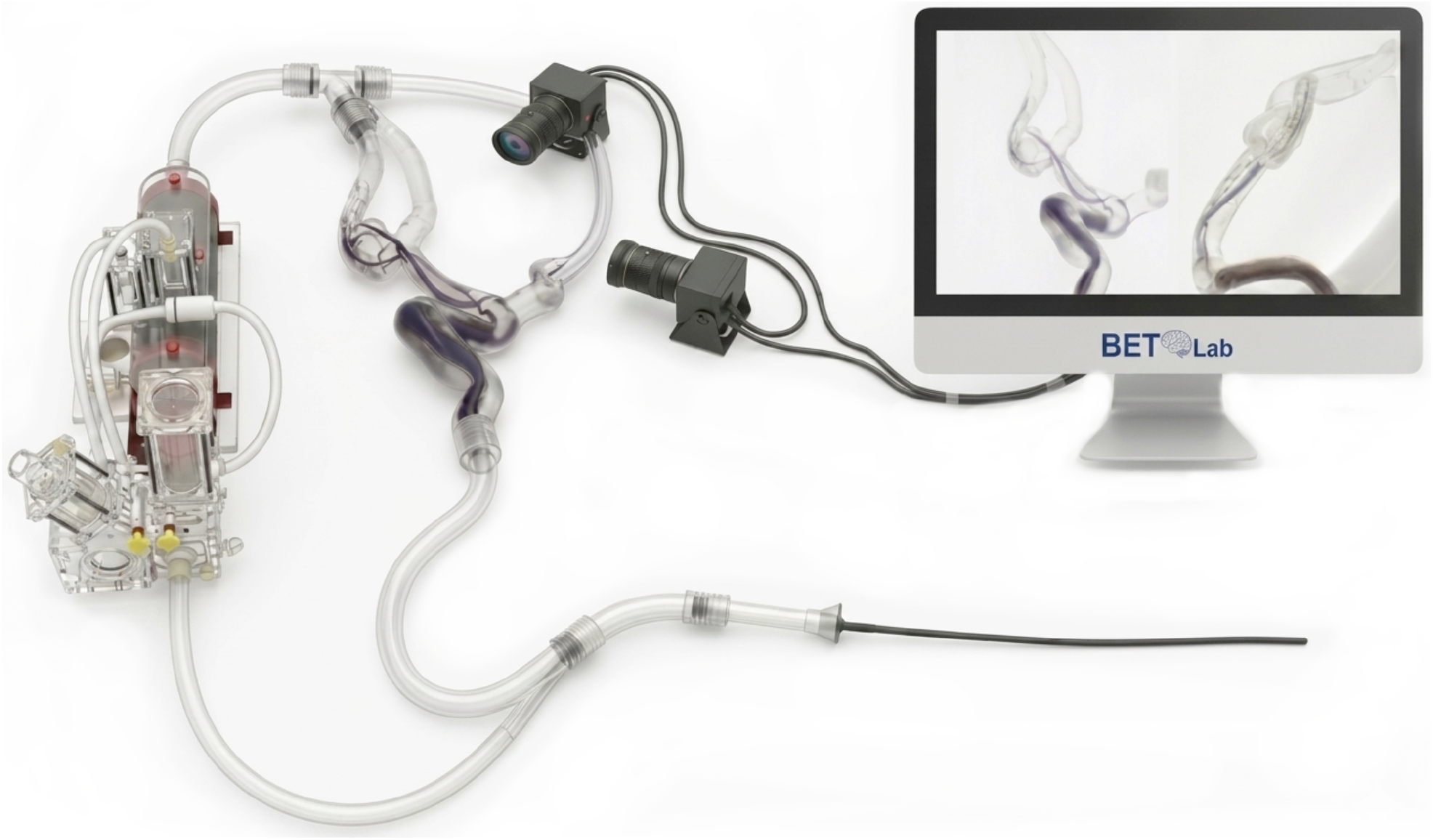
Bench set-up for the pre-procedural testing using a patient-specific model. Patient-specific vascular replicas derived from routine 3D rotational angiography are perfused by a pulsatile pump that reproduces physiological intracranial arterial flow conditions. These replicas are produced in our academic research and development laboratory using computational modeling and additive manufacturing. They are designed to replicate the physical characteristics of the intracranial arteries that are relevant to the behavior of endovascular devices and aneurysm treatment, including the wall friction coefficient, vessel anatomy and the rigidity-to-flexibility ratio. Full optical transparency allows for direct visualization of devices and their behavior under flow, eliminating the need for fluoroscopic guidance and radiation exposure. A biplane camera system provides real-time optical visualization displayed on a dedicated monitor. The operator uses this feedback to assess device behavior and refine the treatment strategy prior to the procedure, navigating and deploying endovascular devices, including microcatheters, guidewires, coils, balloons, stents, and flow diverters, and reproducing the planned procedural steps as in the actual intervention.*\* Some parts of this illustration were created with the assistance of AI; the final image was reviewed and approved by humans*.

### Pre-procedural training

For patients in the PPT group, a benchtop PPT session was conducted before the intervention using a patient-specific vascular replica derived from routine 3D rotational angiography. Replicas were developed by the Brain Endovascular Therapeutic Research and Development Laboratory (University of Geneva, Switzerland) through computational modeling and additive manufacturing. These were intentionally designed to replicate the physical characteristics of intracranial arteries that are relevant to the behavior of endovascular devices and the treatment of aneurysms. This includes a wall friction coefficient that matches the resistance encountered by catheters and devices, a precise reproduction of vessel anatomy, and a rigidity-to-flexibility ratio that mimics the mechanical constraints experienced during device manipulation. The models were developed to be fully optically transparent, allowing devices to be visualized directly under physiological flow conditions. This provides a clear view of the entire device and its behavior, which is not possible under fluoroscopic guidance. It also avoids additional radiation exposure for the operator during the PPT session. Our institution has registered the manufacturing workflow of the models under the trademark *Flow Twin®*.

During the PPT session, operators simulated the planned procedural steps, including the access strategy (selection of the long sheath, intermediate catheter, microcatheter and guidewire), device selection and sizing (coils, balloon, stent or flow diverter), and catheter and device positioning strategy (Figure 2). This session allowed for the confirmation or adaptation of the planned procedural approach prior to the actual intervention and included adjustments to the therapeutic strategy, device sizing, as well as the microcatheter and guidewire shaping. Following the PPT session, operators switched from balloon-assisted coiling (BAC) to stent-assisted coiling (SAC) as the primary strategy in four cases and from SAC to BAC in five cases. Device sizing was modified in 8/24 coiling cases and 5/16 flow diverter cases. Figure 3 provides two illustrative cases demonstrating how PPT influenced the final therapeutic strategy.

**Figure 2:**
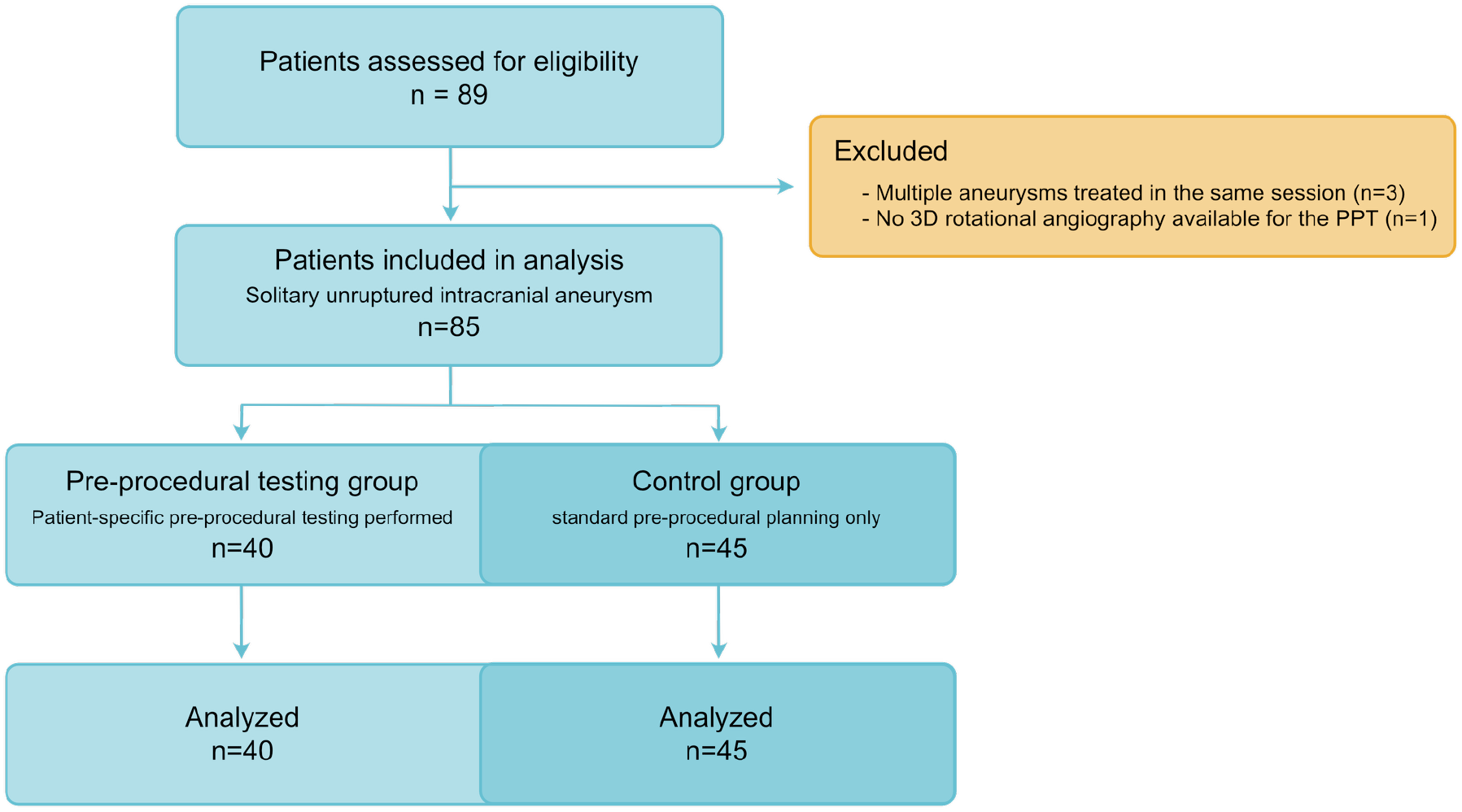
Flow chart of patient selection

**Figure 3:**
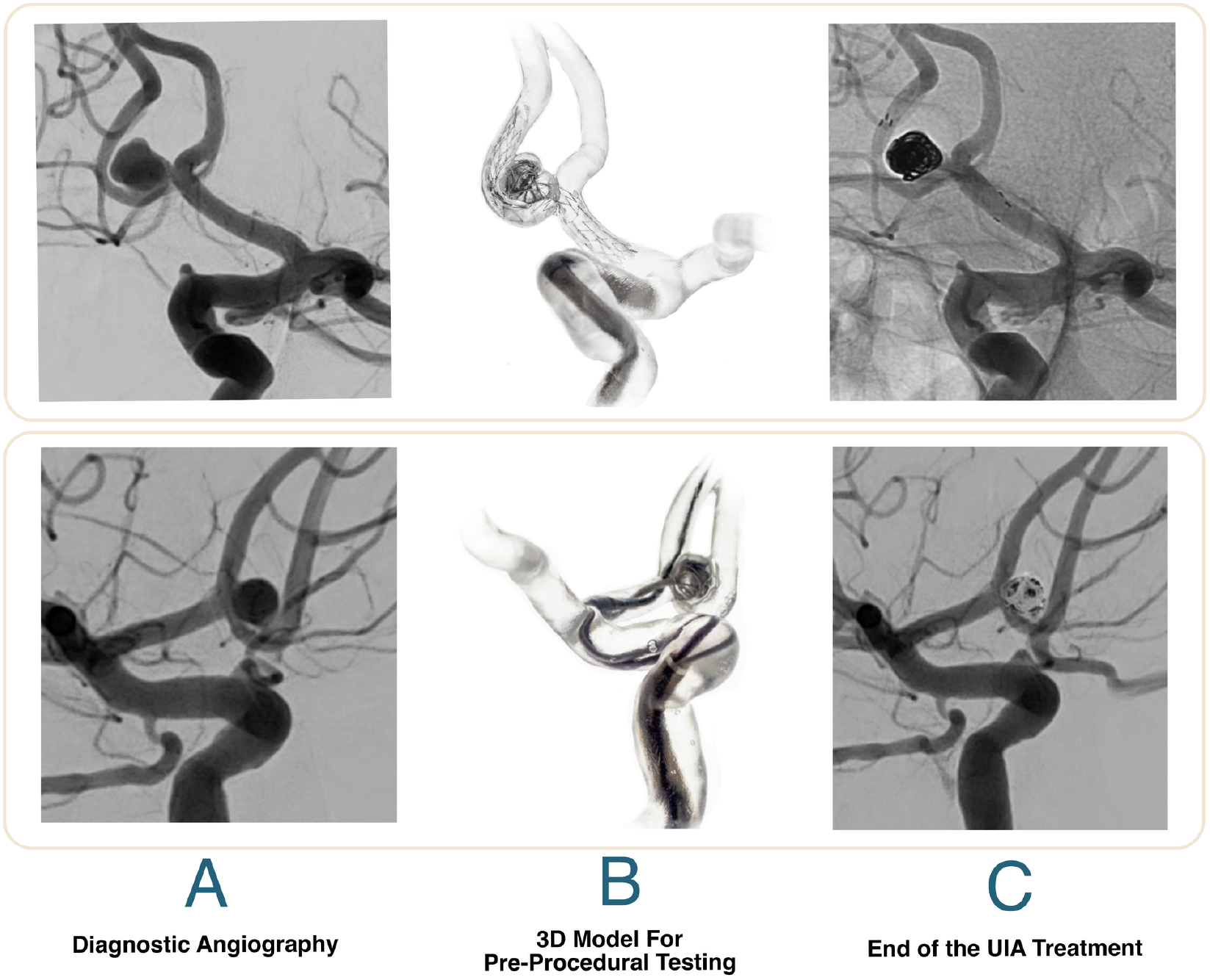
Two illustrative cases of patient-specific pre-procedural testing. (A) Diagnostic cerebral angiography prior to treatment, showing a complex intracranial aneurysm. (B) Pre-procedural testing performed under optical guidance on the patient-specific model of the intracranial arteries and aneurysm. (C) Actual treatment.

After the PPT session and before the procedure, operators completed a structured, prospective evaluation consisting of four items: device behavior realism; model realism; trust in the planned treatment strategy; and perceived utility of the rehearsal. A fifth item, perceived utility after the procedure, was collected immediately after the endovascular treatment. A dedicated research assistant recorded all evaluations using a five-point Likert scale (1 = strongly disagree, 5 = strongly agree).

### Outcomes

#### Primary outcome

This study was designed to primarily assess the feasibility, fidelity and perceived value of patient-specific PPT. Therefore, the primary outcome was defined a priori as an operator-reported composite score reflecting the quality of the rehearsal experience (Table 1). This approach aligns with established frameworks for the feasibility and usability evaluation of medical devices and has been applied in prior neurointerventional and endovascular simulation studies.[18,21,22,24–26] The Training Fidelity Score (TFS) was calculated as the average of three items evaluated immediately following the rehearsal session, ie, device behavior realism; model realism; and confidence in the planned treatment strategy. The internal consistency of the composite score was assessed using Cronbach’s alpha.

**Table 1.**
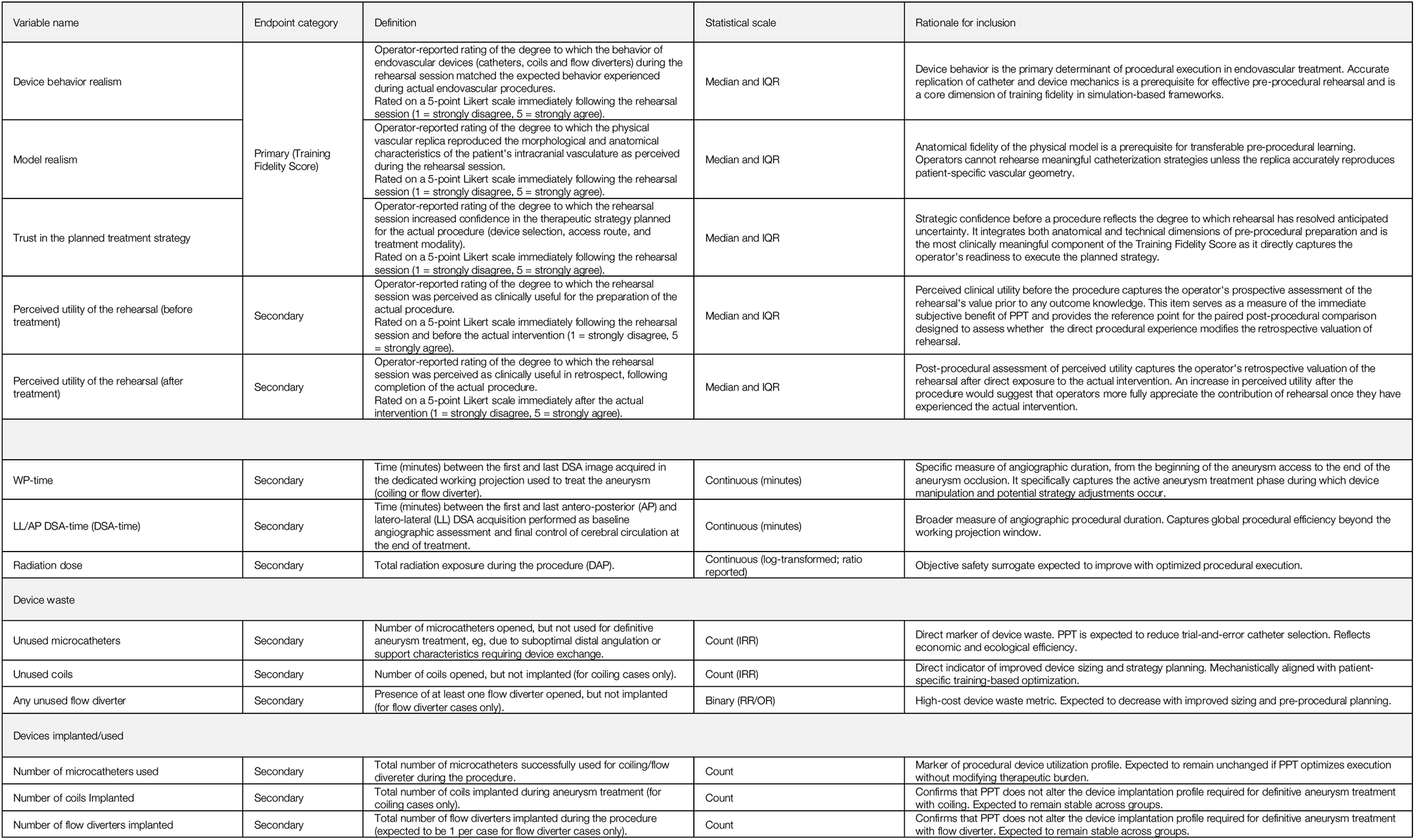
Definition and rationale of study outcomes.

#### Secondary outcomes

Whether operators perceived PPT as useful was assessed using perceived clinical utility, rated immediately after the rehearsal session and again immediately after the actual procedure. This paired assessment was designed to determine if direct experience with the intervention modified the retrospective evaluation of PPT.

Whether PPT stabilizes intraoperative decision-making was assessed by recording unplanned strategy changes during the actual procedure. An intraoperative strategy change was defined as any modification of the therapeutic modality, including conversion from coiling or BAC to flow diversion, or introduction of SAC not anticipated in the pre-procedural plan. The research assistant recorded this variable as a binary outcome immediately following each procedure.

Whether PPT translates into measurable improvements in procedural efficiency was assessed using three objective metrics. Working projection time (WP-time) was defined as the interval between the first and last digital subtraction angiography (DSA) image acquired in the dedicated working projection used to treat the aneurysm. DSA-time was defined as the interval between the first baseline anteroposterior (AP) and latero-lateral (LL) DSA performed after access to the parent artery and the final control DSA obtained at the end of treatment. Radiation exposure was measured as the dose-area product (DAP, Gy·cm^2^) recorded automatically by the angiography system.

Whether PPT reduces unnecessary device use was assessed by comparing device waste and implantation profiles between groups. Device waste was defined as any device that was opened, but not ultimately implanted or used. It was assessed separately for microcatheters, coils and flow diverters. The number of devices ultimately implanted was recorded in parallel to confirm that reductions in waste did not reflect changes in the treatment itself.

Whether procedural efficiency gains were achieved without compromising patient safety was assessed by recording adverse events in both groups. Adverse events leading to transient (<3 months) or permanent morbidity or mortality were recorded according to our predefined institutional criteria.[27]

#### Statistical analysis

All statistical inferential analyses compared the pre-procedural testing (PPT) group with the control group. Since treatment modality (coiling versus flow diversion) is strongly associated with procedure duration and resource use, analyses that included the full cohort were adjusted for treatment modality a priori. The primary adjusted models included treatment modality (coiling versus flow diversion) as a pre-specified covariate. Given the sample size, additional covariates were not included in the primary analysis to prevent model overfitting and maintain interpretability.

Operator-reported Likert scores were summarized using medians and interquartile ranges. The full range of responses for each item was also reported. Cronbach’s alpha coefficient was used to assess the internal consistency of the Training Fidelity Score. A Wilcoxon signed-rank test was used for a paired comparison of perceived clinical utility before and after the procedure.

Continuous outcomes (WP-time, DSA-time and radiation dose) were analyzed using linear regression models that included treatment modality (coil versus flow diverter) as a prespecified covariate. For radiation exposure, the primary analysis used a log-transformed model and the group effect was reported as a ratio of geometric means with 95% confidence intervals (CIs). The effects for WP-time and DSA-time were reported as adjusted mean differences with 95% CIs. Distributions were assessed visually using histograms and Q-Q plots.

Device utilization and waste outcomes were analyzed as counts using generalized linear models with a log link. Poisson regression was used when overdispersion was not present and negative binomial regression was used when overdispersion was present, as determined by the Pearson chi-square statistic divided by degrees of freedom. The results are reported as incidence rate ratios (IRRs) with 95% CIs.

For rare binary outcomes with small cell counts, such as the presence of any unused flow diverter or intraoperative strategy changes, inference was based on Fisher’s exact test. The effect size was expressed as a risk ratio, accounting for zero-cell counts using the Haldane-Anscombe continuity correction.

To assess the reliability of the primary and key secondary continuous outcomes, we conducted prespecified sensitivity analyses for WP-time, DSA-time and radiation dose. These analyses used the same covariate structure as the main analyses. The analyses included trimmed analyses after excluding extreme outliers as defined by Tukey’s rule, Huber M-estimation robust regression and median quantile regression.

We performed the analyses using Python (statsmodels 0.14.4, scipy 1.13.1, and numpy 1.26.4). Statistical significance was set at a two-sided alpha of 0.05. Model assumptions were assessed by inspecting residual distributions and dispersion statistics. No major violations were identified.

#### Sensitivity analysis

We performed prespecified sensitivity analyses for WP-time, DSA-time and radiation dose to evaluate the reliability of the continuous outcomes. These analyses used the same covariate structure as the main analyses, which adjusted for the group effect and treatment modality (coiling versus flow diversion).

1. We examined the influence of extreme outliers using trimmed ANCOVA. We identified extreme outliers for each continuous outcome using Tukey’s rule with wide fences (±3×IQR). Values below Q1 − 3×IQR or above Q3 + 3×IQR were excluded. The adjusted group effect was then re-estimated using the same ANCOVA model on the trimmed dataset.
2. Distributional assumption for radiation dose (log-transformed ANCOVA): as the radiation dose may be right-skewed, we repeated the adjusted analysis of radiation dose using a log-transformed model. We reported the group effect as a ratio of geometric means (PPT versus control) in both the full dataset and after excluding extreme outliers.
3. Robust regression for procedural times: to reduce sensitivity to non-normal residuals and potential leverage points, we repeated the adjusted analyses for WP-time and DSA-time using robust regression with Huber-M estimation (RLM) and median quantile regression. Both were adjusted for treatment modality.

## Results

Among 89 patients assessed for study eligibility, a total of 85 patients met the inclusion criteria: 40 in the PPT group and 45 in the control group. Patient selection is detailed in the supplemental material. Table 2 summarizes the baseline characteristics, which were comparable between groups. Table 3 summarizes the primary and secondary outcomes.

**Table 2.** Patient characteristics.

| Variable | Overall (N=85) | PPT (N=40) | Control (N=45) | p-value |
| --- | --- | --- | --- | --- |
| Age (years) | 56.7 ± 11.1 | 55.9 ± 11.8 | 57.4 ± 10.4 | 0.533 |
| Sex |  |  |  |  |
| Female | 61 (71.8%) | 32 (80.0%) | 29 (64.4%) | 0.139 |
| Male | 24 (28.2%) | 8 (20.0%) | 16 (35.6%) |  |
| Location of the intracranial aneurysm |  |  |  |  |
| A1 | 1 (1.2%) | 0 (0.0%) | 1 (2.2%) | 0.895 |
| AChoA | 2 (2.4%) | 1 (2.5%) | 1 (2.2%) |  |
| ACoA | 20 (23.5%) | 10 (25.0%) | 10 (22.2%) |  |
| Basilar tip | 7 (8.2%) | 2 (5.0%) | 5 (11.1%) |  |
| CarotidT | 5 (5.9%) | 2 (5.0%) | 3 (6.7%) |  |
| MCA | 1 (1.2%) | 1 (2.5%) | 0 (0.0%) |  |
| Ophthalmic | 22 (25.9%) | 12 (30.0%) | 10 (22.2%) |  |
| PCA | 2 (2.4%) | 1 (2.5%) | 1 (2.2%) |  |
| PCom | 16 (18.8%) | 6 (15.0%) | 10 (22.2%) |  |
| PICA | 2 (2.4%) | 1 (2.5%) | 1 (2.2%) |  |
| Pericallosal | 1 (1.2%) | 0 (0.0%) | 1 (2.2%) |  |
| SCA | 4 (4.7%) | 3 (7.5%) | 1 (2.2%) |  |
| V4 | 2 (2.4%) | 1 (2.5%) | 1 (2.2%) |  |
| Aneurysm size (mm) | 7.1 ± 2.2 | 7.0 ± 2.3 | 7.1 ± 2.2 | 0.788 |
| Neck size (mm) | 3.8 ± 1.3 | 3.7 ± 1.1 | 3.9 ± 1.4 | 0.843 |
| Treatment type |  |  |  |  |
| Coiling | 55 (64.7%) | 24 (60.0%) | 31 (68.9%) | 0.530 |
| Flow diverter | 30 (35.3%) | 16 (40.0%) | 14 (31.1%) |  |
| Aneurysm size (mm) (Coiling only) | 6.6 ± 2.1 | 6.2 ± 1.7 | 6.9 ± 2.4 | 0.405 |
| Neck size (mm) (Coiling only) | 3.3 ± 0.9 | 3.1 ± 0.7 | 3.3 ± 0.9 | 0.335 |
| Aneurysm size (mm) (flow diverter only) | 7.9 ± 2.2 | 8.3 ± 2.5 | 7.5 ± 1.6 | 0.851 |
| Neck size (mm) (flow diverter only) | 4.8 ± 1.3 | 4.7 ± 1.0 | 5.0 ± 1.6 | 0.442 |

*Abbreviations:* A1: A1 segment of the anterior cerebral artery; AChoA: anterior choroidal artery; ACoA: anterior communicating artery; BasilarTip: basilar artery tip; CarotidT: carotid artery T-junction (internal carotid artery terminus); MCA: middle cerebral artery; Ophthalmic: ophthalmic artery; PCA: posterior cerebral artery; PCom: posterior communicating artery; PICA: posterior inferior cerebellar artery; Pericallosal: pericallosal artery; PPT: pre-procedural testing, SCA: superior cerebellar artery; V4: V4 segment of the vertebral artery.

**Table 3.**
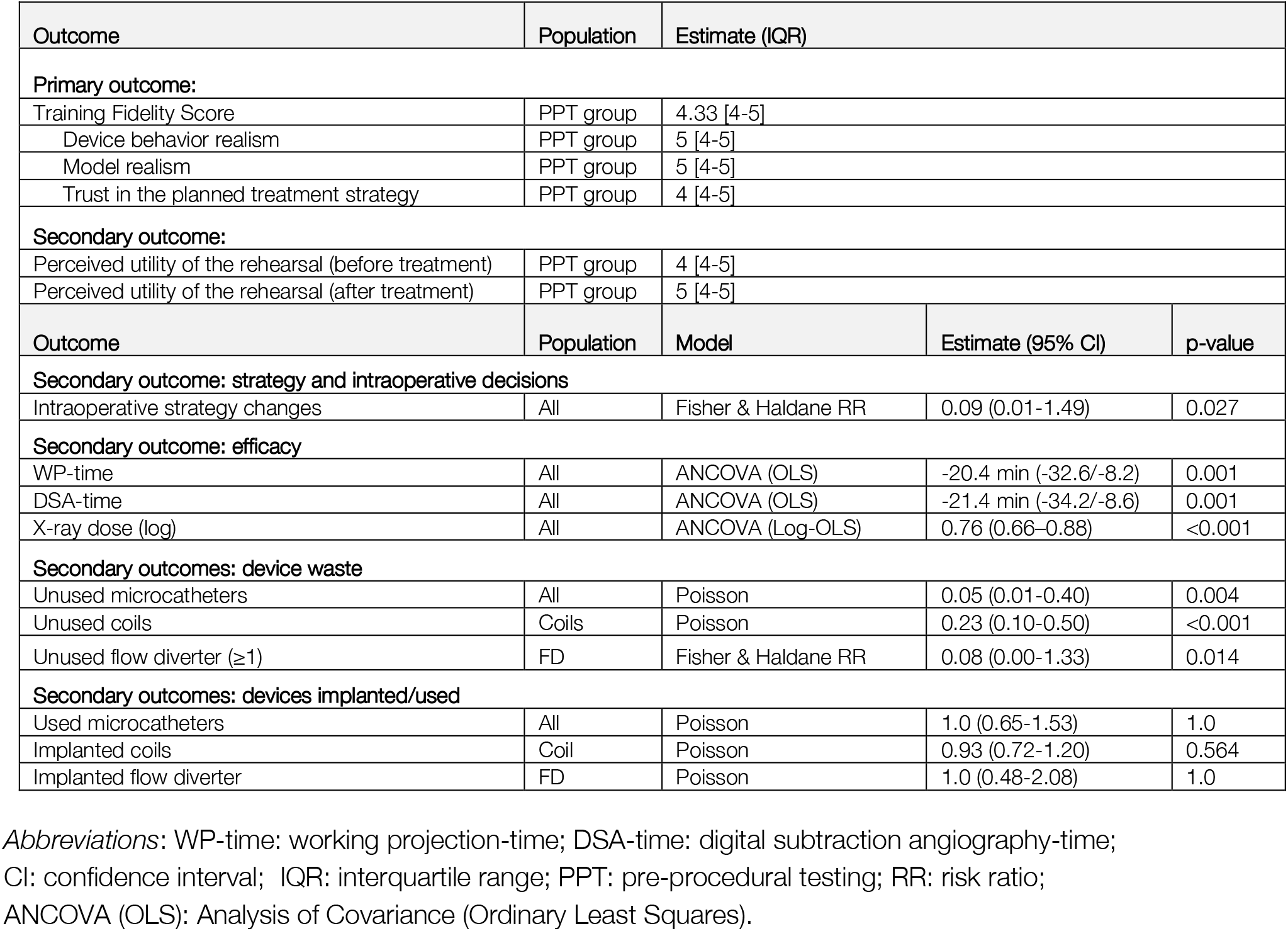
Outcomes.

| Outcome | Population | Estimate (IQR) |  |  |
| --- | --- | --- | --- | --- |
| Primary outcome: |  |  |  |  |
| Training Fidelity Score | PPT group | 4.33 [4-5] |  |  |
| Device behavior realism | PPT group | 5 [4-5] |  |  |
| Model realism | PPT group | 5 [4-5] |  |  |
| Trust in the planned treatment strategy | PPT group | 4 [4-5] |  |  |
| Secondary outcome: |  |  |  |  |
| Perceived utility of the rehearsal (before treatment) | PPT group | 4 [4-5] |  |  |
| Perceived utility of the rehearsal (after treatment) | PPT group | 5 [4-5] |  |  |
| Outcome | Population | Model | Estimate (95% CI) | p-value |
| Secondary outcome: strategy and intraoperative decisions |  |  |  |  |
| Intraoperative strategy changes | All | Fisher & Haldane RR | 0.09 (0.01-1.49) | 0.027 |
| Secondary outcome: efficacy |  |  |  |  |
| WP-time | All | ANCOVA (OLS) | -20.4 min (-32.6/-8.2) | 0.001 |
| DSA-time | All | ANCOVA (OLS) | -21.4 min (-34.2/-8.6) | 0.001 |
| X-ray dose (log) | All | ANCOVA (Log-OLS) | 0.76 (0.66–0.88) | <0.001 |
| Secondary outcomes: device waste |  |  |  |  |
| Unused microcatheters | All | Poisson | 0.05 (0.01-0.40) | 0.004 |
| Unused coils | Coils | Poisson | 0.23 (0.10-0.50) | <0.001 |
| Unused flow diverter (≥1) | FD | Fisher & Haldane RR | 0.08 (0.00-1.33) | 0.014 |
| Secondary outcomes: devices implanted/used |  |  |  |  |
| Used microcatheters | All | Poisson | 1.0 (0.65-1.53) | 1.0 |
| Implanted coils | Coil | Poisson | 0.93 (0.72-1.20) | 0.564 |
| Implanted flow diverter | FD | Poisson | 1.0 (0.48-2.08) | 1.0 |

*Abbreviations:* WP-time: working projection-time; DSA-time: digital subtraction angiography-time; CI: confidence interval; IQR: interquartile range; PPT: pre-procedural testing; RR: risk ratio; ANCOVA (OLS): Analysis of Covariance (Ordinary Least Squares).

### Primary outcome: TFS

The TFS was high across the PPT cohort, indicating that operators consistently perceived the rehearsal as realistic and confidence-building. The median composite score was 4.33/5 (interquartile range [IQR] 4.00–5.00), and 31/40 training sessions (77.5%) achieved a score of 4 or above. The internal consistency of the three-item composite was adequate, with a Cronbach’s alpha of 0.790.

Scores were high across all three individual items. Device behavior realism was rated with a median of 5.0 and 24/40 (60%) training sessions reached the maximum score. PPT realism was rated similarly, with a median of 5 and 23/40 (57.5%) training sessions reached the maximum score. Trust in the planned treatment strategy yielded a median score of 4. The maximum score was reached in 14/40 (35.0%) training sessions and 16/40 (40.0%) sessions reached a score of 4. Figure 4 illustrates the distribution of responses for all items.

**Figure 4:**
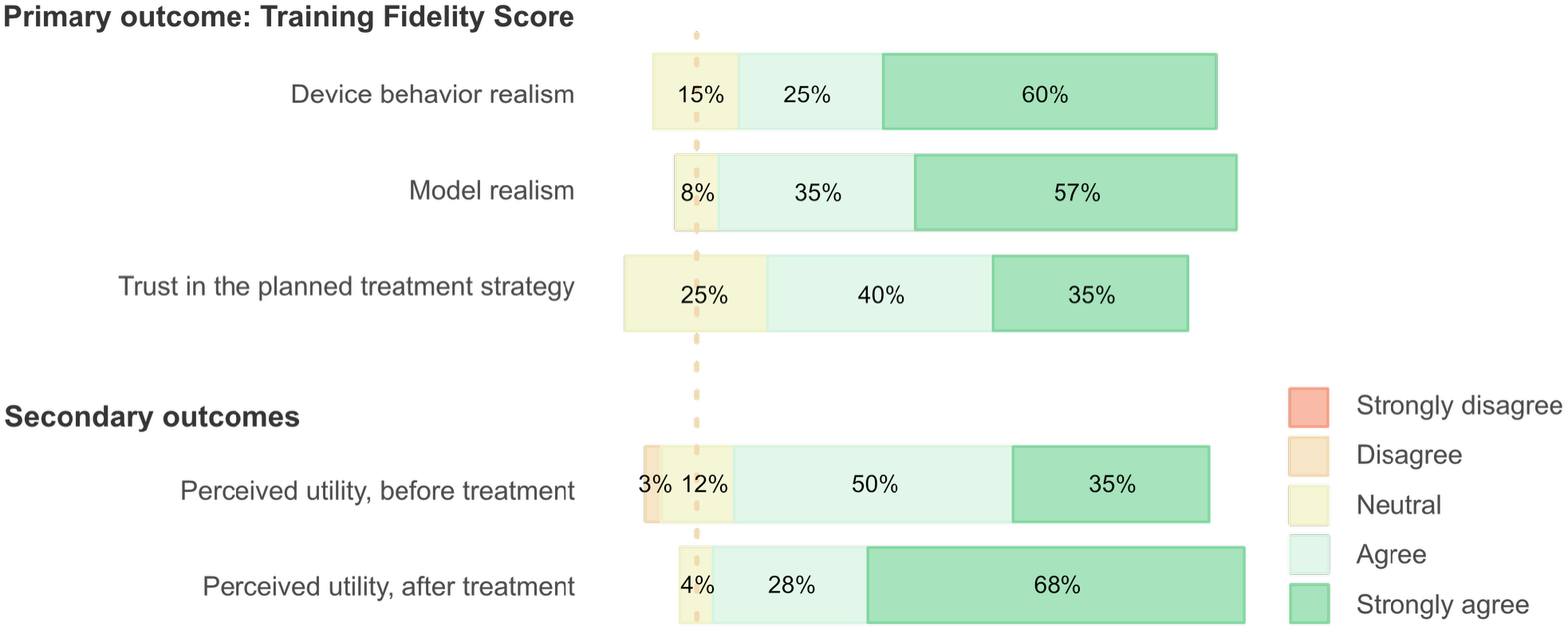
Operator-reported experience after pre-procedural testing. This diverging stacked bar chart shows the distribution of responses to 5 Likert scale items (1 = strongly disagree to 5 = strongly agree) collected from operators after patient-specific pre-procedural testing on the *Flow Twin®*. The upper panel shows the three items that make up the primary outcome, the Training Fidelity Score: device behavior realism; model realism; and trust in the planned treatment strategy. The lower panel displays two secondary outcome items assessing the perceived utility of the rehearsal before and after the actual procedure. Each bar is centered at the neutral response (score 3), with positive responses extending to the right and negative responses extending to the left. Median scores and interquartile ranges are reported for each item.

### Secondary outcomes

#### Operator experience

The perceived clinical utility of the PPT was high before the procedure and increased significantly after the operators experienced the actual treatment. Before the procedure, the median perceived clinical utility was 4, with 14/40 (35%) training sessions reaching the maximum score. Following the procedure, the median score increased to 5, with 27/40 (67.5%) sessions reaching the maximum score. A paired comparison revealed a statistically significant increase in perceived utility following the procedure (Wilcoxon W = 9.5; p=0.0001). Improvement was demonstrated in 18/40 training sessions, while 21 remained unchanged and one showed a decline.

#### Strategy and intraoperative decisions

Pre-therapeutic training was associated with a significant reduction in unplanned intraoperative strategy changes in the PPT group. No intraoperative strategy changes were recorded after any of the 40 PPT sessions, compared to 6/45 changes (13.3%; p=0.027) in the control group. Of the changes observed in the control group, one involved converting from a planned flow diverter to a SAC, two involved converting from a BAC to a flow diverter, and three involved converting from coiling to a SAC. The total number of SAC procedures was comparable between the two groups, with four cases in each, indicating that the strategy changes in the control group reflected intraoperative decision adjustments, rather than a systematic difference in treatment complexity.

#### Procedural efficiency

The PPT group experienced significant reductions in procedural time and radiation exposure. WP-time was reduced by 20.4 min in the PPT group compared to the control group (p=0.001), which corresponds to a 38% relative reduction. DSA-time decreased by 21.4 min (p=0.001), which corresponds to a 33% relative reduction. PPT was also associated with lower radiation exposure (p<0.001), corresponding to a 25% reduction in DAP. Figure 5 illustrates the adjusted marginal means for WP-time and DSA-time.

**Figure 5:**
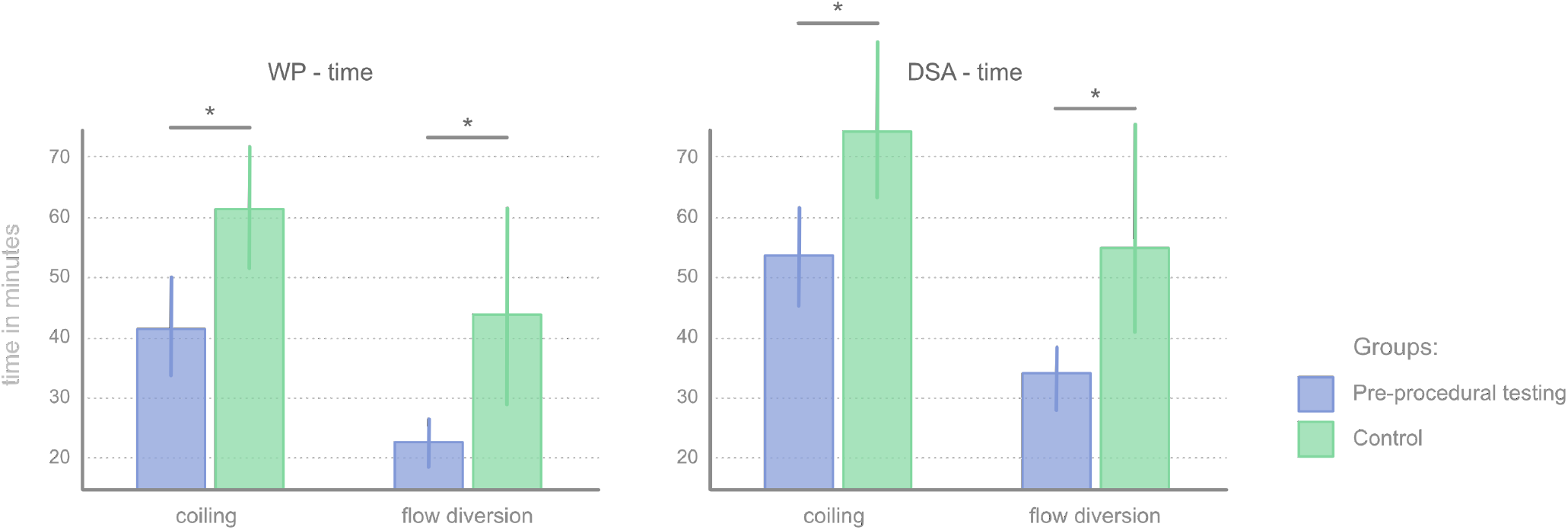
Improved procedural efficiency after pre-procedural testing. Adjusted marginal means and 95% confidence intervals for WP-time (left) and DSA-time (right), estimated from ANCOVA models adjusted for treatment modality. The PPT group demonstrated a 38% reduction in WP-time (adjusted difference −20.4 min; p=0.001) and a 33% reduction in DSA-time (adjusted difference −21.4 min; p=0.001) compared with controls. * p<0.05 *Abbreviations*: WP-time: working projection-time; DSA-time: digital subtraction angiography-time; ANCOVA: Analysis of Covariance; PPT: pre-procedural testing.

**Figure 6:**
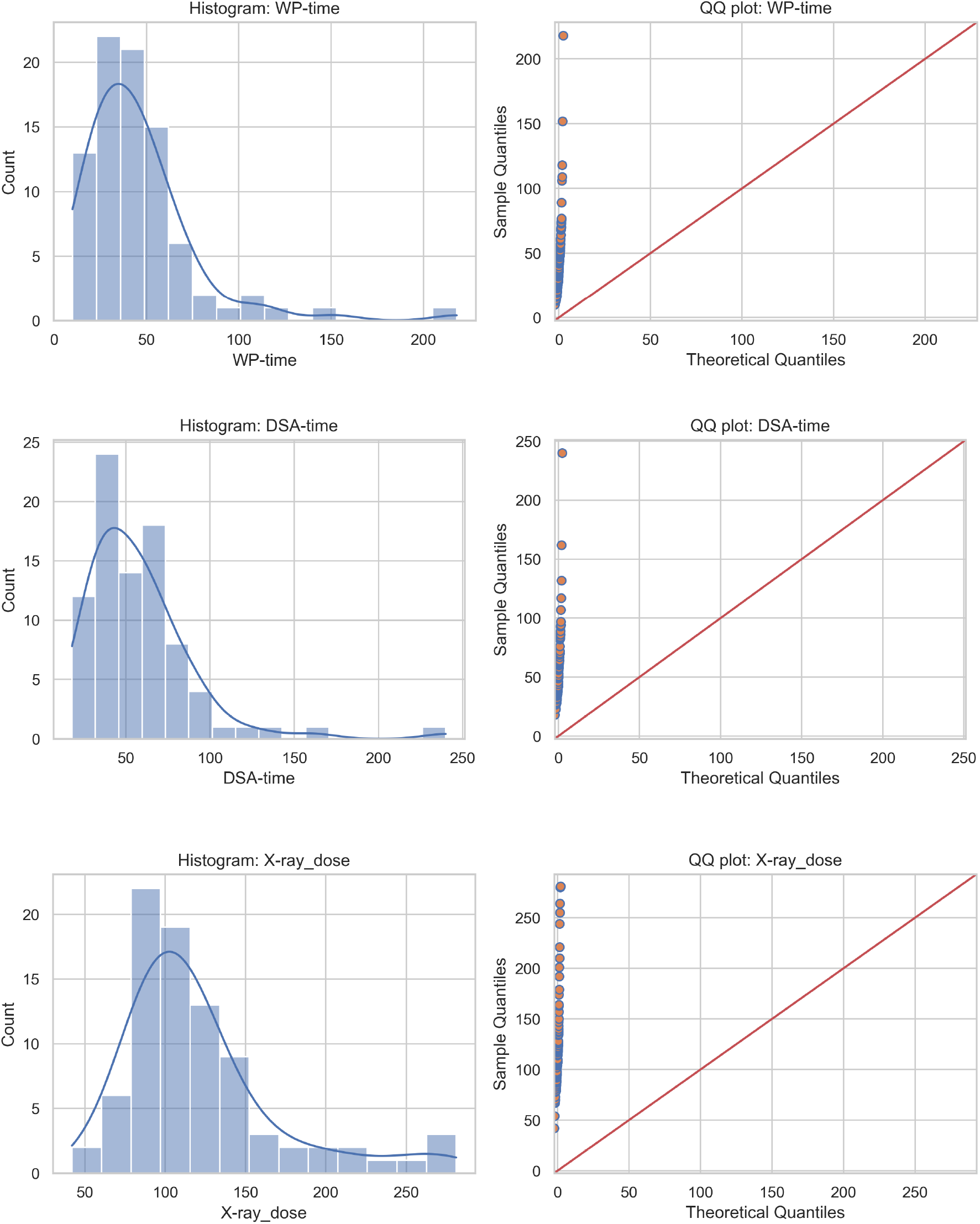
Continuous variable distribution

#### Resource utilization

Device waste decreased significantly across all categories in the PPT group. The incidence of unused microcatheters was significantly lower in the PPT group (p=0.004). Similarly, unused coils were reduced (p<0.001). The occurrence of any unused flow diverters was also lower (p=0.014). However, the number of microcatheters (p=1.0), coils (p=0.564) and flow diverters (p=1.0) used did not differ significantly between groups.

#### Safety

PPT did not compromise procedural safety. Transient morbidity occurred in 3/40 (7.5%) patients in the PPT group and 4/45 (8.9%) in the control group, with no significant difference between the two groups (p=1.00). No permanent morbidity or mortality was observed in either group.

#### Sensitivity analyses

Sensitivity analyses excluding extreme outliers and employing robust and median regression approaches produced consistent results across all continuous outcomes, indicating the robustness of the findings (Table 4):

- WP-time: PPT was associated with a shorter adjusted WP-time in the full ANCOVA model (−20.4 minutes, 95% CI: −32.6 to −8.2, p = 0.001) and after trimming extreme outliers (−14.1 minutes, 95% CI: −22.2 to −6.0, p < 0.001). Robust regression and median quantile regression yielded consistent estimates (Huber: −15.3 min, 95% CI −22.5 to −8.0; median quantile: −14.0 min, 95% CI −23.5 to −4.5; p = 0.004).
- DSA-time: PPT was associated with a shorter adjusted DSA time in the full ANCOVA model (21.4 minutes, 95% CI: −34.2 to −8.6, p = 0.001) and after trimming extreme outliers (14.9 minutes, 95% CI: −23.5 to −6.4, p < 0.001). Robust regression showed a consistent decrease (Huber: −16.7 min, 95% CI: −25.1 to −8.3). Median quantile regression showed a smaller effect (11.0 min, 95% CI −21.5 to −0.6; p = 0.039).
- Radiation dose (DAP): PPT was associated with lower exposure in the full ANCOVA model (−37.2 Gy·cm^2^, 95% CI: −57.1 to −17.2; p <0.001) and after trimming extreme outliers (−30.4 Gy·cm^2^, 95% CI: −48.2 to −12.5; p=0.001). In log-transformed models, PPT was associated with a lower geometric mean dose in the full cohort (ratio: 0.76; 95% CI: 0.66–0.88; p <0.001) and after trimming (ratio: 0.79; 95% CI: 0.69–0.91; p=0.001), corresponding to a 21% to 25% reduction.

**Table 4.** Sensitivity analysis.

| Outcome | Analysis | Estimate (95% CI) | p-value |
| --- | --- | --- | --- |
| WP-time | ANCOVA_full | -20.4 (-32.6/-8.2) | 0.001 |
| WP-time | ANCOVA_trimmed_extreme | -14.1 (-22.2/-6.0) | <0.001 |
| WP-time | Huber | -15.3 (-22.5/-8.0) |  |
| WP-time | Quantile_median | -14.0 (-23.5/-4.5) | 0.004 |
| DSA-time | ANCOVA_full | -21.4 (-34.2/-8.6) | 0.001 |
| DSA-time | ANCOVA_trimmed_extreme | -14.9 (-23.5/-6.4) | <0.001 |
| DSA-time | Huber | -16.7 (-25.1/-8.3) |  |
| DSA-time | Quantile_median | -11.0 (-21.5/-0.6) | 0.039 |
| Radiation dose | ANCOVA_full | -37.2 (-57.1/-17.2) | <0.001 |
| Radiation dose | ANCOVA_trimmed_extreme | -30.4 (-48.2/-12.5) | 0.001 |
| Radiation dose (log) | ANCOVA_full_log | 0.76 (0.66/0.88) | <0.001 |
| Radiation dose (log) | ANCOVA_trimmed_extreme_log | 0.79 (0.69/0.91) | 0.001 |

## Discussion

In this single-center cohort study of consecutive patients undergoing elective endovascular treatment for unruptured intracranial aneurysms, PPT using patient-specific models was associated with high operator-reported training fidelity and perceived clinical utility. These subjective findings were reflected by improvements in all objective secondary outcomes, including a significant reduction in intraoperative strategy changes, shorter procedural times, reduced radiation exposure and less device waste. Together, these findings suggest that PPT improves operator preparedness and procedural execution.

The high TFS observed across the PPT group indicates that operators consistently perceived the testing session as a realistic representation of the procedural environment. These results are consistent with earlier studies evaluating neurointerventional simulator, in which operators rated anatomical realism and device behavior highly on structured, Likert-based questionnaires.[21,22,24,25] Among the three composite items of the TFS, trust in the planned treatment strategy before the intervention showed the widest distribution as strategic confidence requires the integration of anatomical, technical and clinical judgement, not just realism. However, 75% of sessions scored 4 or above for this item, suggesting that PPT consistently supported pre-procedural decision-making. Perceived clinical utility increased significantly after the procedure, suggesting that direct experience of the procedure reinforces the value attributed retrospectively to pre-procedural testing. This elevated subjective confidence was corroborated by the objective secondary outcomes, most notably the significant reduction in intraoperative strategy changes, and was further supported by improvements in procedural efficiency and reductions in device waste.

Patient-specific, pre-therapeutic testing may improve procedural execution through two complementary mechanisms. First, testing on a physical vascular replica allows the catheterization strategy, device selection and anatomical understanding to be confirmed or refined before the procedure begins. Beyond mere confirmation, PPT enables operators to actively identify and select the optimal therapeutic strategy, including the most appropriate device sizing, microcatheter shaping and access route before patient exposure. This reduces the need for intraoperative adjustments, as evidenced by the significant reduction in intraoperative strategy changes in the PPT group. It also translates directly into time-savings and reduced device waste during the actual intervention. Previous neurointerventional simulation studies have demonstrated a high degree of concordance between device selection during testing and actual procedural choices, supporting the idea that model-based testing stabilizes pre-procedural planning.[20,21] These effects were reflected in reductions in procedural time and radiation exposure that were consistent with those reported in patient-specific rehearsal studies in the wider endovascular field.[15–18]

Second, PPT may reduce intraoperative cognitive load by providing operators with a stabilized mental representation of the anticipated procedure, based on direct experience of the behavior of all devices used during the intervention. This schema allows for a more efficient allocation of resources during the live procedure and limits the iterative trial-and-error process that would otherwise be required to identify the optimal approach.[11–13] This reduction in procedural uncertainty is reflected in decreased device waste. Reducing unnecessary device openings has direct economic and environmental implications, as flow diverters, coils and dedicated microcatheters are high-cost, single-use items and opening them unnecessarily generates avoidable expenditure and waste. These efficiency gains were observed in procedures performed exclusively by attending neurointerventionalists with systematic, collegial, pre-procedural planning. This suggests that PPT provides an additional benefit, even for experienced operators working within an optimized standard of care.

The results reported here were obtained using models developed as part of an academic laboratory’s research and development activities (*Flow Twin®*) that were engineered to explicitly reproduce the morphological, physical and biomechanical characteristics of intracranial arteries, aneurysm and endovascular devices. These models were developed for visualization under optical guidance to enable the observation of fine details of the procedure that are inaccessible via conventional fluoroscopy. Although other models based on different technologies are commercially available for neurointerventional simulation, mainly under fluoroscopic guidance, their ability to accurately reflect intracranial arterial mechanics and device behavior has not been formally characterized. Current limitations of PPT include the need for a specialized manufacturing infrastructure and access to endovascular devices for training purposes, both of which are not universally available. Although these barriers are not insurmountable, given that relevant technologies are continually evolving, PPT incurs costs in terms of model fabrication and operator time. However, this seems to be a worthwhile investment rather than an unnecessary expense. Indeed, reductions in procedural time, radiation exposure and device waste suggest a favorable value across the three dimensions relevant to healthcare stakeholders, ie, patient safety, practitioner performance, and payer resource utilization. Integrating PPT as a structured component of pre-interventional preparation with dedicated reimbursement frameworks may represent a next step towards more efficient, reproducible neurointerventional care.

## Conclusions

Patient-specific PPT using vascular replicas was associated with high operator-reported training fidelity, a significant reduction in intraoperative strategy changes, and improvements in procedural efficiency and device waste in elective endovascular treatment of unruptured intracranial aneurysms without compromising safety. These findings support the integration of model-based PPT as a structured component of pre-interventional preparation and provide the basis for a prospective, multicenter validation in the context of endovascular treatment of unruptured intracranial aneurysms.

## Data Availability

All data produced in the present study are available upon reasonable request to the authors.

